# Modulation of inhibitory synaptic plasticity for restoration of basal ganglia dynamics in Parkinson’s disease

**DOI:** 10.1101/2024.08.09.24311371

**Authors:** KA Spencer, A Boogers, S Sumarac, DJ Crompton, LA Steiner, L Zivkovic, Y Buren, AM Lozano, SK Kalia, WD Hutchison, A Fasano, L Milosevic

## Abstract

**Introduction:** Parkinson’s disease is characterized, in part, by hypoactivity of both direct pathway inhibitory projections from striatum to the globus pallidus internus (GPi) and indirect pathway inhibitory projections from globus pallidus externus (GPe) to the subthalamic nucleus (STN), giving rise to disrupted basal ganglia circuit activity. In this study, we explored the use of intracranial stimulation for eliciting long-term potentiation (LTP) of each of these pathologically underactive inhibitory projections for the restoration of basal ganglia circuit dynamics and amelioration of motor symptoms.

**Methods:** Data were collected from a total of 31 people with Parkinson’s disease (42 hemispheres). During deep brain stimulation (DBS) surgery, we assessed microelectrode stimulation-induced changes to inhibitory evoked field potentials (fEP) and hand kinematics before versus after a 40-second train of high-frequency stimulation (HFS) in the GPi (n = 7, 11 sites at 100 Hz) and STN (n = 10, 14 sites at 100 Hz; n = 4, 7 sites at 180 Hz). Additionally, we assessed changes to beta oscillations and hand kinematics in people with chronic DBS implants in the GPi (n = 6 at 125 Hz) and STN (n = 4 at 180 Hz).

**Results:** Intraoperatively, increases in fEP amplitude (p = 0.002) and improved motor performance (p = 0.003) were observed after 100 Hz HFS in the GPi; while in STN, HFS did not potentiate fEPs (p = 0.589) or improve motor performance (p = 0.460) (similar results yielded for 180 Hz in STN). Similarly, extraoperative GPi-DBS produced suppression of beta power (p=0.096) and motor improvement (p = 0.077) before versus after HFS at 125 Hz; while STN-DBS at 180 Hz did not significantly affect beta power (p = 0.267) or motor performance (p=0.850).

**Interpretation:** Our findings support that LTP-like effects in GPi may produce motor improvements that extend beyond stimulation cessation, aligning with optogenetic studies showing long-lasting motor recovery through periodic D1-striatal activation in rodents. The lack of effects in STN suggests that stimulation paradigms may require optimization for effective LTP induction. These findings nevertheless highlight the potential of LTP-based strategies for sustained therapeutic benefits in Parkinson’s disease, warranting further investigation into periodic stimulation paradigms for optimizing DBS efficacy and side effect profiles.

## Introduction

Parkinson’s disease is a complex and progressive neurodegenerative syndrome characterized predominantly by the presence of hypokinetic motor symptoms. The basal ganglia circuit model of posits^1,2^ that degeneration of dopaminergic neurons in the substantia nigra pars compacta (SNc) leads to (i) hypoactivity of D1-mediated striatal “direct pathway” projections to the globus pallidus internus (GPi) and substantia nigra pars reticulata (SNr), as well as (ii) hyperactivity of D2-mediated striatal “indirect pathway” projections to the globus pallidus externus (GPe), which leads to disinhibition of the subthalamic nucleus (STN), contributing further to the overexcitation of basal ganglia output structures (GPi and SNr). Cumulatively, these direct and indirect pathway changes result in excessive inhibition of thalamocortical motor networks, which is thought to give rise to the hypokinetic features of Parkinson’s disease.^2,3^

Deep brain stimulation (DBS) is an established neurocircuit intervention^4^ for the management of the motor symptoms of Parkinson’s disease, which involves the delivery of continuous high frequency stimulation (HFS; ∼130 Hz) to the STN^5^ or GPi.^6^ However, despite its efficacy, continuous HFS can in some cases produce motor^7^ and/or non-motor^8^ side-effects, and is inefficient in terms of battery usage. As such, there is a need to explore and implement novel strategies of neuromodulation which limit the overall duration of stimulation delivery, yet produce functionally-relevant changes to the underlying neurocircuitry which are sustained over time. Neurofeedback-driven closed-loop DBS has been a topic of interest^9,10^ which is still under investigation.^11,12^ Here, we propose an alternative strategy. We sought to investigate whether the elicitation of long-term synaptic plasticity can be leveraged for the restoration of more physiological basal ganglia circuit dynamics and the consequent amelioration of motor symptoms. Specifically, we hypothesize that long-term potentiation (LTP) of pathologically downregulated inhibitory (i) striato-GPi (direct pathway) *or* (ii) GPe-STN (indirect pathway) projections will reduce pathological overactivity (Fig. 1) of basal ganglia output neurons, leading to improvement of hypokinetic features that will persist beyond stimulation delivery.

**Figure 1.**
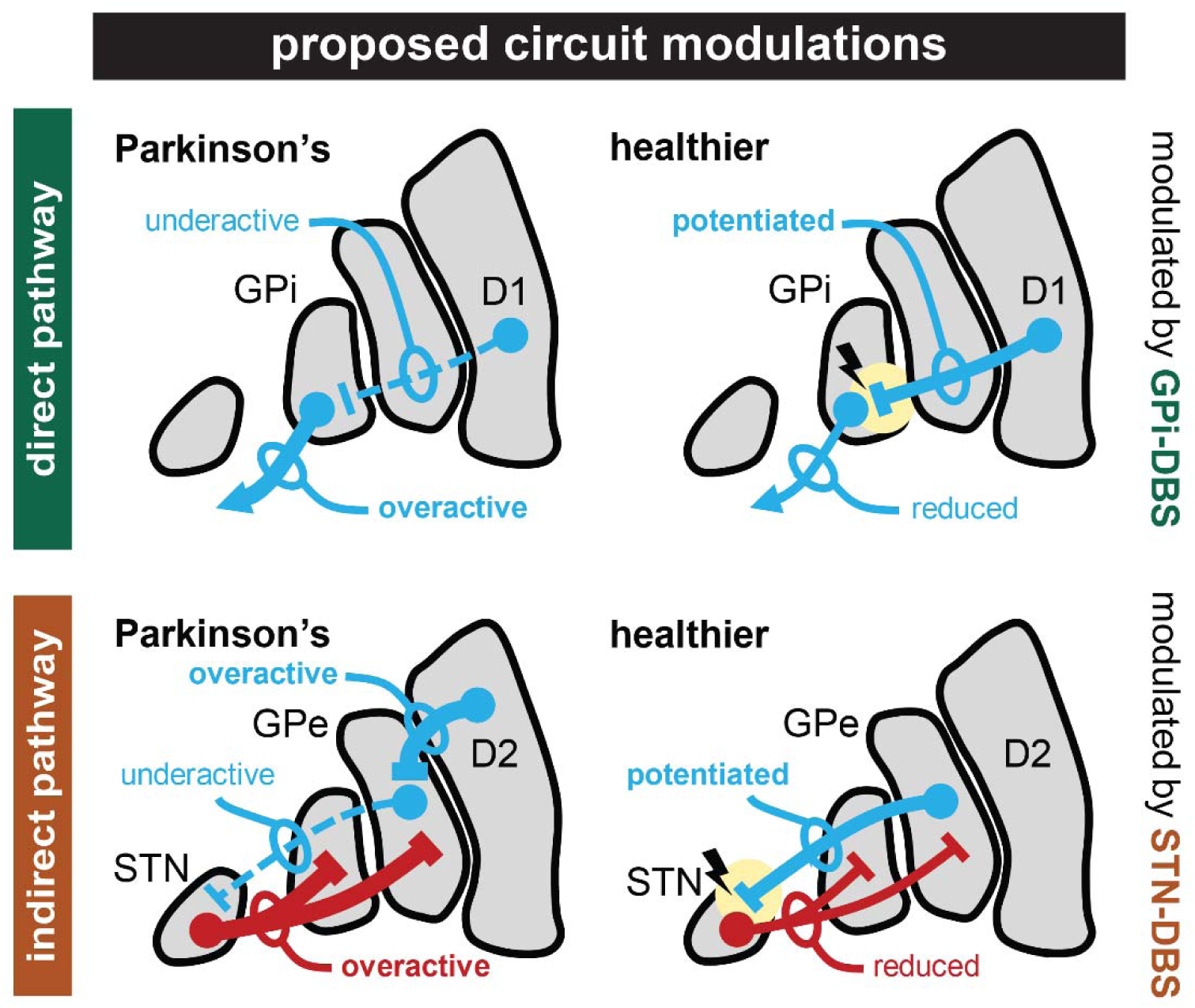
– Potentiation of inhibitory projections within the basal ganglia circuitry. In Parkinson’s disease, inhibitory striato-GPi (top; direct pathway) and GPe-STN (bottom; indirect pathway) projections are underactive. We propose that these fiber pathways can be potentiated by high-frequency GPi- or STN-DBS, respectively, which is expected to lead to improvements in motor symptoms that persist beyond stimulation cessation. GABA = blue; glutamate = red.

While GPi (first interventional target in this work) receives some glutamatergic inputs via STN, the vast majority (∼70%) of afferent innervation is GABAergic, mainly via the striatum.^13–15^ Our previous intraoperative studies have demonstrated that single pulses of electrical stimulation to the GPi produce positive-going evoked field potentials which are coupled with transient periods of neuronal inhibition^16,17^ (i.e., interpreted as stimulus-evoked extracellular GABAergic net responses^18,19^). We have previously demonstrated in patients with Parkinson’s disease that the delivery of a short-duration train of 100 Hz microelectrode stimulation (using a total electrical energy level comparable to clinical DBS)^20^ produced LTP-like responses of these inhibitory inputs to the SNr^21^ and GPi,^16^ confirmed based on the following changes which persisted for several minutes following the cessation of stimulation: increased amplitudes of inhibitory evoked field potentials; prolongation of stimulus-locked neuronal inhibitory silent periods; and a reduction in spontaneous neuronal firing rates. Of relevance, an optogenetic study by the Gittis group showed in dopamine depleted mice that periodic activations of D1 medium spiny neurons (D1-MSNs; i.e., direct pathway striatal neurons) resulted in long lasting motor recovery;^22^ providing support for our hypothesis that potentiating these direct pathway projections may produce benefit which outlasts stimulation (Fig. 1; top). The major source of afferent innervation to STN (second interventional target) is also GABAergic; however, via GPe.^23,24^ Indeed, we have recently described neural substrates of GPe-mediated inhibition to the STN, also in the form of positive-going extracellular inhibitory evoked field potentials (morphologically and functionally distinct from striatum-mediated fields in GPi) which are also correlated with the duration of stimulus-locked neuronal inhibition.^25,26^ Although we have not previously performed LTP experiments in STN, optogenetics studies also from the Gittis group have confirmed that periodic activations of STN-projecting GPe neurons can produce potent long-lasting motor recovery in parkinsonian rodents;^22,27^ providing support for our indirect pathway intervention (Fig. 1; bottom).

## Methods

### Patients

Intraoperative data were collected from patients with Parkinson’s disease undergoing DBS surgery of the GPi (n_GPi_intraoperative_ = 7, unilateral interventions applied at 11 total recording sites) and STN (n_STN_intraoperative_ = 10, 14 recording sites; a supplementary dataset was also collected from an additional 4 patients, 7 recordings sites using a different stimulation frequency as described in detail below). Experiments were conducted at most once per hemisphere. A data summary is available in Supplementary Table 1. Extraoperative data were also collected from patients with Parkinson’s disease at the start of their first DBS programming session at six weeks postoperatively (n_GPi_extraoperative_ = 6; n_STN_extraoperative_ = 4; unilateral experimental interventions performed in one hemisphere only). A data summary is available in Supplementary Table 2. Each patient provided informed consent and the experiments were approved by the University Health Network Research Ethics Board and adhered to the guidelines set by the tri-council Policy on Ethical Conduct for Research Involving Humans.

### General information on intraoperative data acquisition and stimulation

During DBS surgery, two closely-spaced microelectrodes (600 μm spacing; 0.2–0.4 MΩ impedances; ≥10 kHz; sampling rate) were advanced along the surgical trajectory. Recordings were amplified using two Guideline System GS3000 amplifiers (Axon Instruments, USA) and digitized using a CED1401 data acquisition system (Cambridge Electronic Design, UK). All stimulation was active biphasic (cathodal followed by anodal), delivered using an isolated constant current stimulator (Neuro-Amp1A, Axon Instruments). For supplementary intraoperative data in STN, the NeuroPort Signal Processor and CereStim (Blackrock Neurotech, USA) were used for recording and stimulation, respectively. Methods for the electrophysiological identification of the STN^28^ and GPi^29^ were previously published. Briefly, two microelectrodes were advanced in the dorso-ventral direction starting 10 mm above the target. For STN trajectories, STN entry was confirmed by an increase in background noise and recording of single units with firing rates of approximately 20-60 Hz, irregular and bursty firing patterns,^30^ and kinesthetic response to limb movements. Decreases in spiking incidence after 4-6 mm of advancement confirmed exit from the ventral border of the STN. For GPi, recording sites were confirmed based on irregular, high-frequency discharge (60-120 Hz) neurons^31^ with responsiveness to movements. After 10-12 mm of advancement, a decrease in spiking indicated exit from the ventral border, and optic tract was confirmed by visual evoked potentials and stimulation-induced phosphenes.

### Intraoperative experimental paradigm

While within the posteroventromedial GPi (∼2-5 mm from the ventral border) or dorsolateral STN (∼2 mm beyond the dorsal border), patients were asked to perform a motor task involving pronation-supination of their contralateral hand (i.e., asked to perform an action as if screwing in an imaginary lightbulb as fast and as large as possible) for ∼10 seconds while recording accelerometry at baseline (Crossbow Technology, USA). Next, we acquired baseline readouts of inhibitory synaptic efficacy by delivering a set of 10 stimulation pulses at a low frequency (1 Hz; 100 µA; 150 µs width; 10 s) from one microelectrode while recording stimulus-evoked inhibitory field potentials (evoked field potentials; fEPs) using the immediately adjacent microelectrode. Subsequently, a train of stimulation was delivered, intended to elicit LTP (100 Hz; 100 µA; 150 µs width; 40 s), after which low frequency test pulses were delivered again for readouts of post-intervention inhibitory synaptic efficacy and the motor task was repeated. In STN, the supplementary dataset involved delivery of 180 Hz instead of 100 Hz for the tetanizing stimulation.

### Extraoperative experimental paradigm

Extraoperative testing was done during the initial postoperative programming visits in the DBS naïve condition to ensure that no stimulation-related plasticity could previously have occurred. The Percept PC Neurostimulator (Medtronic, USA) was used to record power spectra at all DBS electrode contact pairs,^32^ and the individual contact deduced to have the greatest power of beta frequency (13-30 Hz) oscillations (which happened to be in the left hemisphere for all patients) was selected for unilateral monopolar stimulation as this contact would be expected to produce best clinical result.^33,34^ Stimulation was delivered using a cathodic pulse (passive recharge) of 60 µs pulse width, 2 mA intensity, and 125 Hz for GPi and 180 Hz for STN. A higher frequency was selected for STN-DBS because initial intraoperative results at 100 Hz did not produce significant potentiation of inhibitory evoked field potentials in STN, thus we hypothesized that a higher frequency of tetanic stimulation may be necessary. Patients were asked to perform the pronation-supination task using their right upper limb (incidentally, the more affected side for all patients) prior to stimulation (baseline) while recording data from an Xsens DOT sensor (Movella Technologies, USA) on the index finger. Stimulation was then turned ON for 40 seconds, after which the task was repeated (∼25 s after stimulation cessation) during a 3 min OFF stimulation period; this procedure was repeated three times. Bipolarized local field potential (LFP) data were acquired from contacts surrounding the one used for stimulation using the streaming mode (250 Hz sampling rate) of the Percept PC Neurostimulator.

### Offline analyses and statistics

Intraoperative fEP amplitudes were measured from the pre-stimulus baseline to the peak voltage deflection after each stimulation pulse^21,35^ and normalized relative to pre-stimulation low passed (50 Hz) LFP root mean squared amplitude.^25,26,31^ Extraoperative LFP data were assessed by computing power spectral density (PSD) estimates using Welch’s method (Hanning window; 256 Hz FFT size; data sampled at 250 Hz). PSDs were normalized by dividing by the aperiodic exponent in the 0.5-100 Hz range using the Fitting Oscillations and One-Over-F (FOOOF) model,^36^ and oscillatory power in the beta range was calculated as the summated power within a 4 Hz window around the detected beta peak. For analysis of pronation-supination motor task performance, the intraoperative accelerometer (hardware vector summed and directly calculated as an analog voltage input) and extraoperative IMU sensor (digitally vector summed from X, Y, Z acceleration components) traces were downsampled to 60 Hz using polyphase filtering and 1Hz high pass filtered using a 3rd order Butterworth filter.^37^ Then, movement amplitude was calculated by taking the difference between the average peaks and troughs. All pre- vs post-HFS data (fEP amplitudes, LFP power, and movement amplitudes) were compared using both frequentist and Bayesian two-tailed paired samples t-tests using JASP (version 0.18.3). For extraoperative data, these t-tests were used to compare baseline data versus the first post-HFS epoch; however, repeated-measures ANOVA tests were also used which considered all three trials.

## Data availability

All fEP, LFP, and accelerometry data are provided in a public repository (see: https://osf.io/am2q8/)

## Results

### Intraoperative results

In GPi, we found strong evidence (based on Bayesian statistics) of an effect of HFS on inhibitory fEP amplitudes (p = .002; BF_10_ = 28.116) and on hand pronation-supination movement amplitude (p = .003; BF_10_ = 17.975); however, in STN, we found anecdotal evidence of a lack of effect of HFS on fEP (p = .589; BF_10_ = 0.309) and movement (p = .460; BF_10_ = 0.329) amplitudes (Fig. 2). Further, we found moderate evidence of a difference in fEP change when comparing GPi and STN (p = .007; BF_10_ = 6.641), as well as anecdotal evidence of a difference in movement amplitude change (p = .028; BF_10_ = 2.454) following HFS (not depicted). Of note, movement analyses were performed for all recording sites; however, two recording sites from GPi and one recording site from STN had to be excluded from fEP analyses as there were no discernable fEPs in the recording traces. Supplementary results for STN stimulation at 180 Hz (p=.182, BF10 = 0.795) were similar to STN results at 100 Hz.

**Figure 2.**
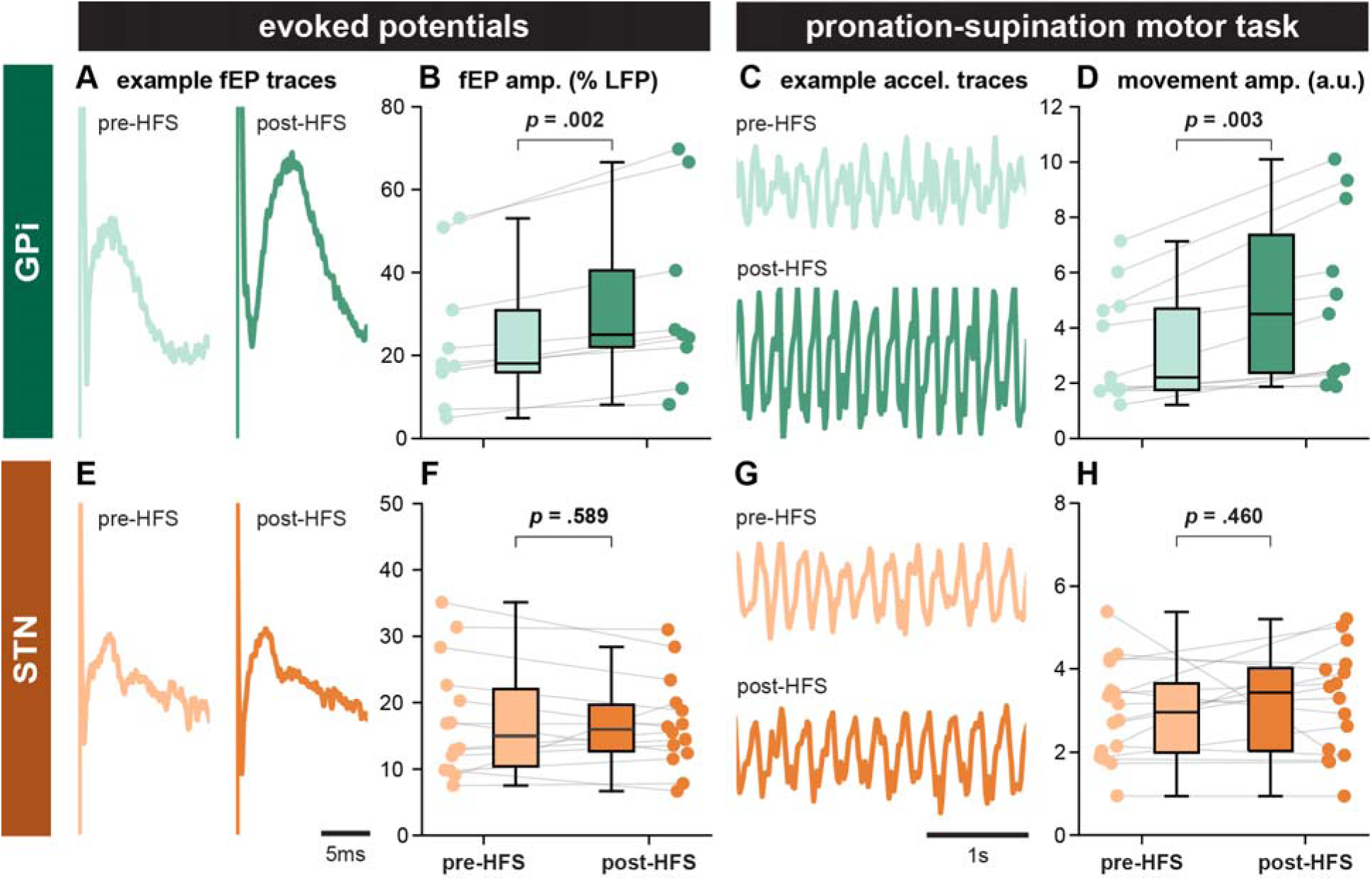
– Intraoperative results. Increases in (A/B) evoked field potential (fEP) amplitude and (C/D) hand pronation-supination amplitude were found after a train of high-frequency microstimulation (40 s; 100 Hz; 100 μA; 150 μs pulse width) in GPi, but not (E/F/G/H) STN.

### Extraoperative results

When comparing baseline data to the first post-HFS epoch in GPi, we found anecdotal evidence of an effect of HFS on beta power (p = 0.096; BF_10_ = 1.312) and on movement amplitude (p = .077; BF_10_ = 1.534) (Fig. 3). In STN, we found anecdotal evidence of a lack of effect of HFS on beta power (p = 0.267; BF_10_ = 0.760) and movement amplitude (p = 0.850; BF_10_ = 0.435) (Fig. 3). Further, we found anecdotal evidence of a difference in beta power change when comparing GPi and STN (p = 0.034; BF_10_ = 2.347), though anecdotal evidence of a lack of difference in movement amplitude change (p = 0.779; BF_10_ = 0.511) following HFS (not depicted). Of note, for the second GPi patient, data from the second trial were used, since for the first trial, the motor task was inadvertently initiated prior to termination of stimulation. ANOVA analyses on LFP and movement data which considered all three post-HFS trials were not significant (p > 0.3; BF_10_ < 1.0) for both GPi and STN, potentially as a result of experimental fatigue.

**Figure 3.**
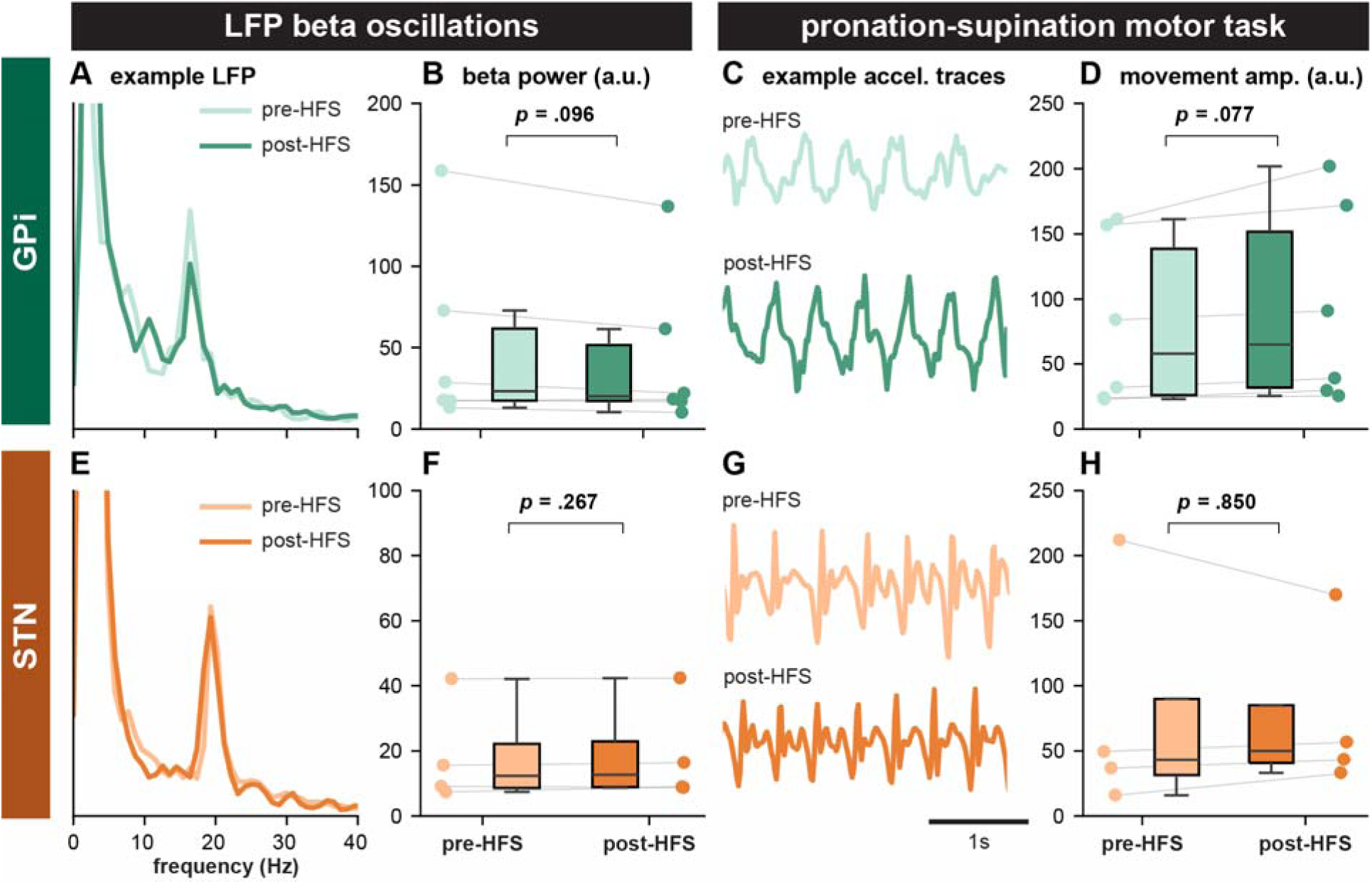
– Extraoperative results. (A/B) Decreases local field potential (LFP) beta power and (C/D) increases hand pronation-supination amplitude were found after a train of high-frequency stimulation (40 s; 125 Hz; 2 mA; 60 µs pulse width) in GPi, but not (E/F/G/H) STN (40 s; 180 Hz; 2 mA; 60 µs pulse width).

## Discussion

### Corroborating animal and human studies

The hypotheses and proposed circuit interventions (Fig. 1) related to this study were largely motivated by an optogenetics work by Mastro and colleages.^22^ In parkinsonian rodents, the authors showed that periodic (ON 40 s / OFF 3 min) activations of (direct pathway) D1-MSNs could produce motor recovery (improved immobility / bradykinesia) that outlasted stimulation cessation. The same study also showed that non-selective activation of GPe neurons could not reproduce this phenomenon; however, cell-type specific (i) activation of parvalbumin (PV) - expressing GPe neurons and/or (ii) suppression of lim homeobox 6 (Lhx6) - expressing GPe neurons could produce long-lasting benefits.

In our intraoperative studies, a 40 s train of microstimulation produced an increase in the amplitude positive-going fEPs representing inhibitory inputs to the GPi, coupled with an increase in the frequency and amplitude of a sequential hand movements in a motor task for assessing hand bradykinesia. These results may therefore corroborate the direct pathway optogenetics results described above;^22^ while additionally suggesting that lasting benefits may be the result of potentiating striato-pallidal projections, which are otherwise downregulated in Parkinson’s disease. On the other hand, our applied stimulation paradigms were not able to produce LTP-like effects to GPe-STN projections (also downregulated in Parkinson’s disease) and no clinical improvements were observed. To this end, the ability to produce LTP-like effects of striato-pallidal but not GPe-STN projections may be the result of differences in functional properties of engaged synapses which represent the major sources of inhibitory inputs to respective targets. Indeed, in recent work, our team has demonstrated differential dynamics of short-term synaptic plasticity of these projections, in that striato-nigral (/pallidal) projections were subject to rapid and potent synaptic depression during 100 Hz stimulation, while GPe-STN projections were able to maintain inhibitory drive with even with a high rate of activation.^25^ Thus, the selective resiliency of GPe-STN synapses with high-frequency stimulation may preclude the necessary signaling cascade to trigger LTP responses. Indeed, we have previously shown that the stimulation frequency threshold for eliciting LTP-like responses of striato-nigral projections coincides with the frequency at which synaptic depression occurs during the stimulation train intended to produce the LTP-like response.^21^ As such, an even higher stimulation frequency in STN may be necessary to produce LTP.

### Mechanisms of inhibitory LTP

In general, far less is known about inhibitory LTP compared to excitatory.^38^ Briefly, it is thought that the process is mediated by trafficking of GABA_A_ receptors (GABA_A_Rs) from intracellular sites to the synaptic membrane.^39^ From rodent slice work, it is thought that the process of inhibitory LTP is also the result of NMDA receptor dependent (which may concurrently be activating as a result of DBS due to its non-selective nature), producing exocytosis and therefore increased surface expression of GABA_A_Rs at the dendritic surface of hippocampal neurons, resulting in increases in the amplitudes of miniature IPSPs.^40^ Trafficking of GABA_A_Rs has been shown to be dependent on GABA_A_R associated protein (GAB_A_RAP), glutamate receptor interacting protein (GRIP), and Ca2+ calmodulin dependent kinase II (CaMKII).^40–42^

### Clinical considerations

In terms of the temporal response to stimulation changes in the clinical context, it is well described that tremor is a fast-responding symptom to the cessation of STN-DBS (seconds to minutes), while there is a significantly slower worsening of bradykinesia and rigidity (half an hour to an hour) and axial symptoms (>1 hour).^43^ The clinical phenomenon of a latent return of symptom severity after chronic stimulation may indeed be the result of elicitation of a beneficial form of LTP in the basal ganglia circuit. Trials explicitly comparing the temporal response of symptoms to the onset/offset of stimulation in STN versus GPi are lacking, but it has been reported that pallidal stimulation is associated with longer washout periods after deactivation of stimulation^44^ (historically considered “not important clinically because patients are unlikely to deactivate their stimulation systems”^45^). Indeed, our own team’s clinical experience in GPi-DBS for Parkinson’s disease indicates a similar latency for rigidity, but more variable and delayed effects for signs such as bradykinesia, and dystonia/dyskinesia. Accordingly, switching STN-DBS off in most cases leads to quicker return of motor signs whereas what is observed in GPi-DBS is more variable and usually delayed with the exception of rigidity. Additional knowledge on the comparative effects of kinetics of changes to STN and GPi stimulation comes from studies in patients with dystonia, where there is convincing evidence that dystonia is faster responding to STN-DBS than GPi-DBS.^46^ With regards to DBS for dystonia, it has also been reported that symptoms can take up to two days to return after cessation of stimulation, indeed attributed to long-term neural reorganization and plasticity in the motor system.^47,48^

### Future directions

While our results for STN-DBS remain inconclusive, our work may nevertheless represent a translation of circuit manipulations derived in optogenetics studies to the human neuromodulation space. With regards to GPi-DBS, it is possible that periodic stimulation may be able to produce sufficient clinical benefit while greatly reducing the overall amount of stimulation, which has the possibility to reduce side effects associated with continuous HFS and delay battery replacement surgeries (if/when relevant). In our pilot studies in the extraoperative environment, we opted for a higher frequency of activation for STN-DBS, under the hypothesis that a higher rate of activation may produce LTP. Indeed, more recent translational work from the Gittis group has in fact shown that there may exist an optimized combination of extracellular stimulation frequency (175 Hz) and duration (200 ms) that can be applied in a bursting manner (at 1 Hz intervals), which optimally mimics optogenetics manipulations of PV-GPe neuronal upregulation / Lhx6-GPe neuronal downregulation. As such, further refinement of the stimulation paradigm in the human context may indeed produce better results (e.g., even higher stimulation frequencies than attempted here). Overall, periodic stimulation, already in use as a conventional approach in DBS for epilepsy,^49^ may have to potential to improve current DBS applications for Parkinson’s disease and may also serve as a framework for the development of novel approaches for other intractable neurological conditions which are thus far inadequately managed. Further studies are warranted to establish translatability / long-term efficacy of this approach.

### Limitations

Within this work, the applied intraoperative stimulation (100uA, 150us pulse width, assuming 0.2 MΩ impedance) was comparable to a clinical DBS intensity of 2 mA (assuming 130 Hz, 60 µs pulse width, and 1 kΩ impedance)^20^ in terms of total electrical energy delivered^50^ (∼30 µJ/s); though of a different current density.^51^ Indeed, 2 mA clinical stimulation is expected to produce benefit, while the intraoperative stimulation parameters used here have also been shown to be able to produce potent tremor suppression.^52^ Other limitations of this work include the relatively small sample sizes, inability to assess the time course of LTP-like phenomena due to time constraints, and the inability to use pharmacological agents to verify synapse/cell-type specificity. Moreover, DBS a non-selective intervention expected to activate all afferent projects in the vicinity of stimulation. To this end, extracellular stimulation is expected to produce compound effects, likely representing summated activity of activated pathways.^53,54^ While net inhibitory effects on spike firing^16,25,26^ suggest the predominance of GABAergic synaptic input activations in both STN and GPi,^18,19^ this does not rule out co-activation of other inputs. Importantly, both GPe and striatal GABAergic inputs converge in GPi; however, GPe inputs only represent ∼15% of the innervation compared to ∼70% represented by striatum.^15^ Indeed, what are thought to be GPe-mediated responses have been shown to be able to be elicited by DBS in GPi;^55^ however, given that GPe-mediated response in STN were not subject to LTP (shown in the work herein), the same would be expected of these projections to GPi if inadvertently co-activated by our GPi intervention. To this end, another limitation of this work is that our interventions targeting GPe-STN projections were generally inconclusive (even when exploring higher stimulation frequencies) in terms of producing changes to neural activity or motor performance; however, these results may serve as an affirmative negative control, in that the lack of LTP-like results is indeed not expected to produce benefit that outlasts stimulation cessation.^22^ On this note, the fact that STN-DBS did not produce prolonged suppression of beta is perhaps surprising and contradictory to early reports.^56^ However, more recent studies indeed have shown very transient post-stimulation suppression of beta activity^57^ as well as near-instantaneous returns to baseline following stimulation cessation.^58^ Indeed, it is suggested that the after-effects may depend on the duration of HFS, supported by findings that 300 s of stimulation produced lasting attenuation of beta activity while 30s did not.^59^

### Conclusion

In the intraoperative environment, we found that a 40 s train of high frequency microstimulation (100 Hz) in the GPi produced LTP-like effects of inhibitory striato-pallidal projections, coupled with improved performance on a clinical motor task evaluating contralateral hand bradykinesia which persisted beyond stimulation delivery. However, high frequency microstimulation (100 or 180 Hz) in STN did not produce LTP-like effects of inhibitory GPe-STN projections, nor improved motor performance. In the extraoperative environment, we found that 40 s blocks of GPi-DBS (125 Hz) led to suppression of beta frequency LFP oscillations and improved motor performance which outlasted stimulation delivery; whereas results for STN-DBS (180 Hz) remained variable / inconclusive. As such, the GPi results, particularly in GPi, suggest that long-lasting therapeutic benefit may be able to be achieved by eliciting LTP of pathologically underactive basal ganglia pathways; and may therefore pave the way for new-generation periodic stimulation paradigms.

## Data Availability

All data produced are available online at https://osf.io/am2q8/

## Acknowledgements

The authors would like to thank the participants for their contributions to the work.

## References

1. DeLong, M. R. Primate models of movement disorders of basal ganglia origin. Trends Neurosci. 13, 281–285 (1990).

2. McGregor, M. M. & Nelson, A. B. Circuit Mechanisms of Parkinson’s Disease. Neuron 101, 1042–1056 (2019).

3. Bezard, E., Brotchie, J. M. & Gross, C. E. Pathophysiology of levodopa-induced dyskinesia: potential for new therapies. Nat. Rev. Neurosci. 2, 577–588 (2001).

4. Neumann, W.-J., Horn, A. & Kühn, A. A. Insights and opportunities for deep brain stimulation as a brain circuit intervention. Trends Neurosci. (2023) doi:10.1016/j.tins.2023.03.009.

5. Kumar, R. et al. Double-blind evaluation of subthalamic nucleus deep brain stimulation in advanced Parkinson’s disease. Neurology 51, 850 (1998).

6. Kumar, R. et al. Deep brain stimulation of the globus pallidus pars interna in advanced Parkinson’s disease. Neurology 55, S34–9 (2000).

7. Odekerken, V. J. et al. Subthalamic nucleus versus globus pallidus bilateral deep brain stimulation for advanced Parkinson’s disease (NSTAPS study): a randomised controlled trial. Lancet Neurol. 12, 37–44 (2013).

8. Funkiewiez, A. et al. Long term effects of bilateral subthalamic nucleus stimulation on cognitive function, mood, and behaviour in Parkinson’s disease. J. Neurol. Neurosurg. Psychiatry 75, 834–839 (2004).

9. Priori, A., Foffani, G., Rossi, L. & Marceglia, S. Adaptive deep brain stimulation (aDBS) controlled by local field potential oscillations. Exp. Neurol. 245, 77–86 (2013).

10. Little, S. et al. Adaptive deep brain stimulation in advanced Parkinson disease. Ann. Neurol. 74, 449–457 (2013).

11. Iturrate, I. et al. Beta-driven closed-loop deep brain stimulation can compromise human motor behavior in Parkinson’s Disease. bioRxiv 696385 (2019) doi:10.1101/696385.

12. Little, S. & Brown, P. Debugging Adaptive Deep Brain Stimulation for Parkinson’s Disease. Mov. Disord. 35, 555–561 (2020).

13. Parent, A. & Hazrati, L.-N. Functional anatomy of the basal ganglia. II. The place of subthalamic nucleus and external pallidium in basal ganglia circuitry. Brain Res. Rev. 20, 128–154 (1995).

14. Bolam, J. P., Hanley, J. J., Booth, P. a. C. & Bevan, M. D. Synaptic organisation of the basal ganglia. J. Anat. 196, 527–542 (2000).

15. Shink, E. & Smith, Y. Differential synaptic innervation of neurons in the internal and external segments of the globus pallidus by the GABA- and glutamate-containing terminals in the squirrel monkey. J. Comp. Neurol. 358, 119–141 (1995).

16. Milosevic, L. et al. Modulation of inhibitory plasticity in basal ganglia output nuclei of patients with Parkinson’s disease. Neurobiol. Dis. 124, 46–56 (2019).

17. Liu, L. D. et al. Frequency-dependent effects of electrical stimulation in the globus pallidus of dystonia patients. J. Neurophysiol. 108, 5–17 (2012).

18. Yoshida, M. & Precht, W. Monosynaptic inhibition of neurons of the substantia nigra by caudatonigral fibers. Brain Res. 32, 225–228 (1971).

19. Precht, W. & Yoshida, M. Blockage of caudate-evoked inhibition of neurons in the substantia nigra by picrotoxin. Brain Res. 32, 229–233 (1971).

20. Milosevic, L. et al. Subthalamic suppression defines therapeutic threshold of deep brain stimulation in Parkinson’s disease. J. Neurol. Neurosurg. Psychiatry 90, 1105–1108 (2019).

21. Milosevic, L. et al. Neuronal inhibition and synaptic plasticity of basal ganglia neurons in Parkinson’s disease. Brain 141, 177–190 (2018).

22. Mastro, K. J. et al. Cell-specific pallidal intervention induces long-lasting motor recovery in dopamine-depleted mice. Nat. Neurosci. 20, 815–823 (2017).

23. Baufreton, J. et al. Sparse but Selective and Potent Synaptic Transmission From the Globus Pallidus to the Subthalamic Nucleus. J. Neurophysiol. 102, 532–545 (2009).

24. Kita, T. & Kita, H. The Subthalamic Nucleus Is One of Multiple Innervation Sites for Long-Range Corticofugal Axons: A Single-Axon Tracing Study in the Rat. J. Neurosci. 32, 5990– 5999 (2012).

25. Steiner, L. A. et al. Persistent synaptic inhibition of the subthalamic nucleus by high frequency stimulation. Brain Stimulat. 15, 1223–1232 (2022).

26. Steiner, L. A. et al. Neural signatures of indirect pathway activity during subthalamic stimulation in Parkinson’s disease. Nat. Commun. 15, 3130 (2024).

27. Spix, T. A. et al. Population-specific neuromodulation prolongs therapeutic benefits of deep brain stimulation. Science 374, 201–206 (2021).

28. Hutchison, W. D. et al. Neurophysiological identification of the subthalamic nucleus in surgery for Parkinson’s disease. Ann. Neurol. 44, 622–628 (1998).

29. Hutchison, W. D. et al. Differential neuronal activity in segments of globus pallidus in Parkinson s disease patients: NeuroReport 5, 1533–1537 (1994).

30. Scherer, M., et al. Single-neuron bursts encode pathological oscillations in subcortical nuclei of patients with Parkinson’s disease and essential tremor. Proc. Natl. Acad. Sci. 119, e2205881119 (2022).

31. Sumarac, S. et al. Interrogating basal ganglia circuit function in Parkinson’s disease and dystonia. eLife 12, (2023).

32. Jimenez-Shahed, J. Device profile of the percept PC deep brain stimulation system for the treatment of Parkinson’s disease and related disorders. Expert Rev. Med. Devices 18, 319– 332 (2021).

33. Tinkhauser, G. et al. Directional local field potentials: A tool to optimize deep brain stimulation. Mov. Disord. 33, 159–164 (2018).

34. Milosevic, L. et al. Online Mapping With the Deep Brain Stimulation Lead: A Novel Targeting Tool in Parkinson’s Disease. Mov. Disord. 35, 1574–1586 (2020).

35. Prescott, I. A. et al. Levodopa enhances synaptic plasticity in the substantia nigra pars reticulata of Parkinson’s disease patients. Brain 132, 309–318 (2009).

36. Donoghue, T. et al. Parameterizing neural power spectra into periodic and aperiodic components. Nat. Neurosci. 23, 1655–1665 (2020).

37. Spooner, R. K., Bahners, B. H., Schnitzler, A. & Florin, E. Time-resolved quantification of fine hand movements as a proxy for evaluating bradykinesia-induced motor dysfunction. Sci. Rep. 14, 5340 (2024).

38. Kullmann, D. M., Moreau, A. W., Bakiri, Y. & Nicholson, E. Plasticity of Inhibition. Neuron 75, 951–962 (2012).

39. Luscher, B., Fuchs, T. & Kilpatrick, C. L. GABAA Receptor Trafficking-Mediated Plasticity of Inhibitory Synapses. Neuron 70, 385–409 (2011).

40. Marsden, K. C., Beattie, J. B., Friedenthal, J. & Carroll, R. C. NMDA receptor activation potentiates inhibitory transmission through GABA receptor-associated protein-dependent exocytosis of GABA(A) receptors. J. Neurosci. Off. J. Soc. Neurosci. 27, 14326–14337 (2007).

41. Marsden, K. C., Shemesh, A., Bayer, K. U. & Carroll, R. C. Selective translocation of Ca2+/calmodulin protein kinase IIα (CaMKIIα) to inhibitory synapses. Proc. Natl. Acad. Sci. 107, 20559–20564 (2010).

42. Chen, Z.-W. & Olsen, R. W. GABAA receptor associated proteins: a key factor regulating GABAA receptor function. J. Neurochem. 100, 279–294 (2007).

43. Temperli, P. et al. How do parkinsonian signs return after discontinuation of subthalamic DBS? Neurology 60, 78–81 (2003).

44. Shulman, L. M. et al. The Clinically Important Difference on the Unified Parkinson’s Disease Rating Scale. Arch. Neurol. 67, 64–70 (2010).

45. Follett, K. A. et al. Pallidal versus subthalamic deep-brain stimulation for Parkinson’s disease. N. Engl. J. Med. 362, 2077–2091 (2010).

46. Sun, B., Chen, S., Zhan, S., Le, W. & Krahl, S. E. Subthalamic nucleus stimulation for primary dystonia and tardive dystonia. Acta Neurochir. Suppl. 97, 207–214 (2007).

47. Ruge, D. et al. Shaping reversibility? Long-term deep brain stimulation in dystonia: the relationship between effects on electrophysiology and clinical symptoms. Brain 134, 2106– 2115 (2011).

48. Ruge, D. et al. Deep brain stimulation effects in dystonia: Time course of electrophysiological changes in early treatment. Mov. Disord. 26, 1913–1921 (2011).

49. Salanova, V. et al. The SANTÉ study at 10 years of follow-up: Effectiveness, safety, and sudden unexpected death in epilepsy. Epilepsia 62, 1306–1317 (2021).

50. Koss, A. M., Alterman, R. L., Tagliati, M. & Shils, J. L. Calculating total electrical energy delivered by deep brain stimulation systems. Ann. Neurol. 58, 168–168 (2005).

51. Maggio, F. et al. Micro vs macro electrode DBS stimulation: A dosimetric study. Annu. Int. Conf. IEEE Eng. Med. Biol. Soc. IEEE Eng. Med. Biol. Soc. Annu. Int. Conf. 2010, 2057– 2060 (2010).

52. Milosevic, L. et al. Physiological mechanisms of thalamic ventral intermediate nucleus stimulation for tremor suppression. Brain 141, 2142–2155 (2018).

53. Milosevic, L. et al. A theoretical framework for the site-specific and frequency-dependent neuronal effects of deep brain stimulation. Brain Stimulat. 14, 807–821 (2021).

54. Neumann, W.-J., Steiner, L. A. & Milosevic, L. Neurophysiological mechanisms of deep brain stimulation across spatiotemporal resolutions. Brain 146, 4456–4468 (2023).

55. Steiner, L. A. & Milosevic, L. A convergent subcortical signature to explain the common efficacy of subthalamic and pallidal deep brain stimulation. Brain Commun. 5, fcad033 (2023).

56. Kühn, A. A. et al. High-Frequency Stimulation of the Subthalamic Nucleus Suppresses Oscillatory β Activity in Patients with Parkinson’s Disease in Parallel with Improvement in Motor Performance. J. Neurosci. 28, 6165–6173 (2008).

57. Eusebio, A. et al. Deep brain stimulation can suppress pathological synchronisation in parkinsonian patients. J. Neurol. Neurosurg. Psychiatry 82, 569–573 (2011).

58. Feldmann, L. K. et al. Subthalamic beta band suppression reflects effective neuromodulation in chronic recordings. Eur. J. Neurol. 28, 2372–2377 (2021).

59. Bronte-Stewart, H. et al. The STN beta-band profile in Parkinson’s disease is stationary and shows prolonged attenuation after deep brain stimulation. Exp. Neurol. 215, 20–28 (2009).

