## Supplementary Table 1 for "Modulation of inhibitory synaptic plasticity for restoration of basal ganglia dynamics in Parkinson’s disease"

**Supplementary Table 1: Intraoperative patient data summary**

| **patient ID** | **structure** | **hemisphere** | **behaviour?** | **fEP?** | **stimulation frequency** |
| --- | --- | --- | --- | --- | --- |
| 1 | STN | left | Yes | Yes | 100 |
| 1 | STN | right | Yes | No | 100 |
| 2 | STN | right | Yes | Yes | 100 |
| 3 | STN | left | Yes | Yes | 100 |
| 3 | STN | right | Yes | Yes | 100 |
| 4 | STN | right | Yes | Yes | 100 |
| 4 | STN | left | Yes | Yes | 100 |
| 5 | STN | right | Yes | Yes | 100 |
| 6 | GPi | left | Yes | Yes | 100 |
| 6 | GPi | right | Yes | Yes | 100 |
| 7 | STN | right | Yes | Yes | 100 |
| 7 | STN | left | Yes | Yes | 100 |
| 8 | STN | right | Yes | Yes | 100 |
| 9 | STN | right | Yes | Yes | 100 |
| 9 | STN | left | Yes | Yes | 100 |
| 10 | STN | left | Yes | Yes | 100 |
| 10 | STN | right | Yes | No | 100 |
| 11 | STN | left | Yes | No | 100 |
| 12 | GPi | left | Yes | No | 100 |
| 13 | GPi | left | Yes | Yes | 100 |
| 14 | GPi | left | Yes | No | 100 |
| 15 | GPi | right | Yes | Yes | 100 |
| 15 | GPi | left | Yes | Yes | 100 |
| 16 | GPi | right | Yes | Yes | 100 |
| 16 | GPi | left | Yes | Yes | 100 |
| 17 | GPi | right | Yes | Yes | 100 |
| 17 | GPi | left | Yes | Yes | 100 |
| 18 | STN | right | No | Yes | 180 |
| 18 | STN | left | No | Yes | 180 |
| 19 | STN | left | No | Yes | 180 |
| 20 | STN | right | No | Yes | 180 |
| 20 | STN | left | No | Yes | 180 |
| 21 | STN | left | No | Yes | 180 |
| 21 | STN | right | No | Yes | 180 |

**Note:** The shaded rows in the table represent patients who did not undergo behavioral assessments. Instead, these patients received 180Hz stimulation in the subthalamic nucleus (STN) to investigate potential plasticity effects at higher frequencies. Recordings for these patients were conducted using the NeuroPort Signal Processor and CereStim systems (Blackrock Neurotech, USA). For all other recordings, the Axon system was employed, utilizing Guideline System GS3000 amplifiers (Axon Instruments, USA) for amplification, and digitization was carried out using a CED1401 data acquisition system (Cambridge Electronic Design, UK).
