## Supplementary Table 2 for "Modulation of inhibitory synaptic plasticity for restoration of basal ganglia dynamics in Parkinson’s disease"

**Supplementary Table 2: Extraoperative patient data summary**

| **patient ID** | **structure** | **hemisphere** | **LFP bipolar configuration** | **UPDRSIII (baseline, trial1, trial2, trial3)** |
| --- | --- | --- | --- | --- |
| 1 | STN | left | 1-3 | 3, 3, 3, 3 |
| 2 | STN | left | 1-3 | 3, 3, 2, 3 |
| 3 | GPi | left | 0-2 | 2, 2, 2, 2 |
| 4 | GPi | left | 1-3 | 2, 2, 2, 2 |
| 5 | GPi | right | 1-3 | 2, 2, 2, 2 |
| 6 | GPi | left | 1-3 | 3, 2, 2, 3 |
| 7 | GPi | right | 1-3 | 3, 2, 2, 2 |
| 8 | STN | left | 1-3 | 1, 1, 2, 2 |
| 9 | STN | left | 0-2 | 3, 2, 2, 2 |
| 10 | GPi | right | 1-3 | 1, 1, 2, 2 |
